# Comparison of three sequencing methods for identifying and quantifying antibiotic resistance genes (ARGs) in sewage

**DOI:** 10.1101/2024.04.01.24305146

**Authors:** Divya Mondhe, Farah Ishtiaq

## Abstract

**Background:** Globally, antimicrobial resistance (AMR) poses a critical threat, requiring robust surveillance methodologies to tackle the growing challenge of drug-resistant microbes. AMR is a huge challenge in India due to high disease burden, lack of etiology-based diagnostic tests and over the counter availability of antibiotics and inadequate treatment of wastewaters are important drivers of AMR in India. There is lack of effective surveillance platforms that monitor health-associated infections. This include developing an understanding on background levels of AMR in the environment and comparison of AMR monitoring methods.

**Objectives:** This study evaluated the performance of three AMR sequencing methods, Illumina AmpliSeq AMR panel, QIAseq xHYB AMR panel and shotgun sequencing method for the detection of antimicrobial resistance genes (ARGs) in urban sewage. Our goal is to provide insights into the application and robustness of each sequencing method.

**Methods:** We compare the prevalence, diversity, and composition of ARGs across sequencing method and by sample type (inlet vs outlet) in four sewage treatment plants (STP).

**Results:** Regardless of the sequencing method used the dominant ARGs remained consistent, and their differential analysis showed consistent trends in detection of epidemiologically relevant ARGs. The cost-effectiveness analysis revealed comparable per-sample costs, with amplicon-based sequencing offering specificity for targeted genes, and shotgun sequencing uses a whole-genome sequencing approach that provides high-resolution taxonomic information for the characterisation of pathogens. This methodology can only detect ARGs that have been annotated in the reference database (e.g., CARD). Therefore, some novel types of ARGs present in the samples may be missed since the analysis is based on a similarity search. Differential abundance analysis to understand change in abundance in dominant ARGs from inlet to outlet of STP showed consistent trends across methods. However, with caution raised regarding potential artifacts introduced by enrichment steps in QIAseq xHYB AMR panel.

**Conclusion:** The choice of panel used would be governed by the context of the study. Nonetheless, our exploratory study shows that the data gathered using different sequencing pipelines helps in quantifying the ARG burden in the environment. This information is crucial in understanding the spatio-temporal distribution of ARGs in different environment and could help in developing PCR-based approaches for targeted surveillance.

## 1. Introduction

Since the groundbreaking discovery of penicillin in 1928, the advent of numerous antimicrobial agents has revolutionized the treatment of infectious diseases in humans, plants, and animals [1]. However, the overuse and misuse of these antibiotics have led to the emergence of drug-resistant microbes, undermining the efficacy of conventional treatment strategies. This surge in AMR poses a critical global threat, contributing to increased pathogen persistence, elevated rates of communicable disease transmission, and rendering existing therapeutic interventions ineffective [2]. Recognizing AMR as a leading cause of global mortality, the World Health Organization (WHO) emphasizes the urgency for robust antimicrobial stewardship practices and enhanced surveillance worldwide [3].

The One Health concept emphasizes the interconnectedness of human, animal, and ecosystem health. AMR transmission spans across these three components, with the environment acting as a crucial bridge between them. Effective AMR surveillance is pivotal in defining the scope of the problem, developing interventions, and mitigating selection pressures [4]. Current surveillance methods, which rely on passive reporting of phenotypes in clinical isolates, often present logistical challenges and lack holistic community-level information [4,5]. In contrast, sewage or environmental samples emerge as attractive candidates for community-level surveillance due to their unique role as environmental reservoirs of AMR [6].

Sewage provides ideal environments for the emergence and persistence of AMR, facilitating both horizontal and vertical transmission of ARGs [6]. While several studies have employed sequencing-based approaches for AMR surveillance, the multitude of available sequencing methods adds complexity to the selection process. This study aims to address this challenge by comparing the performance of three distinct AMR sequencing methods: amplicon based AmpliSeq AMR Panel and QIAseq xHYB AMR Panel, and shotgun metagenomic sequencing. Using urban sewage, we aim to provide insights into the robustness, cost effective and applicability of each sequencing method in profiling the ARG composition in sewage samples.

Metagenomic antibiotic resistome data are multi-dimensional, often represented several ARGs in metagenomes, and therefore inferring meaningful results is not only dependent on the abundance of single ARGs, but also on the overall scope of the resistome such as the diversity of associated ARGs [7]. Therefore, the exploration of resistome composition is typically evaluated by alpha diversity (within sample) and beta diversity (between sample) differences in profiling or analysis as it is usually employed for determining differences in microbial diversity [7]. In general, the variation in ARG distribution within and between samples is typically assessed by calculating alpha and beta diversity indices respectively, similar to practices employed for determining differences in microbial community profiling [8,9]. To understand the efficiency of sewage treatment plant in removal of ARGs, differentially abundant genes in metagenomic samples are used. Our analysis explores limitations of sequencing method while drawing inferences in assessing the efficiency of sewage treatment. This comparative analysis aims to contribute to the optimization of ARG surveillance methodologies using environmental samples and priorities for developing targeted ARG monitoring.

## 2. Material and Methods

### 2.1. Sewage sampling

Grab samples from inlet and outlet of four STPs in Bengaluru were collected between March and July 2021 (Table 1). These STP follow activated sludge process for treating the sewage water in inlet (primary filtration unit) and release the treated water in outlet in an effluent tank before releasing the processed water into the environment. Each 80ml sewage sample was collected in sterile plastic bottle which was sealed immediately after collection and transported to the laboratory at 4LJ. Subsequently, the samples were stored at -20LJ until further processing.

**Table 1:** Number of ARGs detected by sequencing method and sample type.

| Sewage Treatment Plant | Collection month | Sample type | AmpliSeq AMR panel | QIAseq xHYB AMR panel | Shotgun Sequencing |
| --- | --- | --- | --- | --- | --- |
| (Capacity in Million litre per day) |  |  | Number of Antimicrobial Resistance Genes |  |  |
| KC1 (218MLD) | April | Inlet | 152 | 701 | 284 |
|  |  | Outlet | 128 | 630 | 216 |
| LBH (1.5 MLD) | March | Inlet | 146 | 669 | 284 |
|  |  | Outlet | 96 | 528 | 119 |
| RCL (40 MLD) | July | Inlet | 132 | 661 | 252 |
|  |  | Outlet | 126 | 625 | 220 |
| YLK (10 MLD) | March | Inlet | 147 | 615 | 240 |
|  |  | Outlet | 136 | 626 | 222 |

### 2.2. Sample processing and DNA extractions

Samples were thawed for 24 hours at 4LJ. The inlet samples were filtered using muslin cloth to remove larger solid particles and debris. Subsequently, vacuum filtration was carried out using a 0.22 µm membrane filter (MF-Millipore® Membrane Filter). These membrane filters were then utilized for DNA extraction using the QIAamp® DNeasy PowerWater Kit (Cat. No. 14900-100-NF) (Qiagen, Germany), following the manufacturer’s instructions. A negative control was included in each extraction batch. The extracted DNA samples were quantified using the Qubit, and their A260/A280 ratios were measured using a nanodrop.

### 2.3. Next-Generation Sequencing for Antimicrobial Resistance Genes (ARG)

Samples were sequenced using the following three sequencing methods, i) AmpliSeq AMR panel, ii) QIAseq xHYB AMR Panel and iii) Shotgun metagenomic sequencing. Simultaneously, 16S rRNA amplicon-based sequencing was performed to assess the bacterial diversity in these samples. The bacterial diversity observed with 16S rRNA sequencing was compared with that of shotgun sequencing.

#### 2.3.1. Illumina AmpliSeq AMR Panel

The AmpliSeq panel has 2 pools of a total of 815 amplicons targeting the presence of 478 AMR genes. The DNA extracts were screened using multiplexed primer pools for the presence of ARGs following manufacturers protocols. Amplicon libraries were prepared using AmpliSeq Library PLUS for Illumina kit (Cat. No. 20019102). The quality of the library was assessed using a 2100 Bioanalyzer with a high sensitivity assay kit (Agilent CA, USA). These libraries were further sequenced using Miseq system with 2 x 150bp paired-end chemistry.

#### 2.3.2. QIAseq xHYB AMR Panel

The QIAseq xHYB AMR panel targets 2786 ARGs. The Amplicon libraries for were prepared using the QIAseq FX DNA Library kit (Cat. No. 180477). The quality of these libraries was assessed using a 2100 Bioanalyzer with a high sensitivity assay kit (Agilent CA, USA). The libraries were pooled for xHYB-target enrichment and sequenced on NovaSeq 600 system with 2 x 100bp paired-end chemistry.

#### 2.3.3. Shotgun sequencing

The shotgun libraries were prepared using Illumina® DNA Prep kit, (M) Tagmentation (Cat. No. 20018705). The quality of the library was assessed using a 2100 Bioanalyzer with a high sensitivity assay kit (Agilent CA, USA) and were further sequenced on NovaSeq 600 system with 2 x 100bp paired-end chemistry.

#### 2.3.4. 16S RNA amplicon-based sequencing

The V3-V4 hypervariable regions of bacterial 16S rRNA genes in the genomic DNA were PCR amplified using the forward primer 338F (5’ CTACGGGNGGCWGCAG) and reverse primer 802R (5’ CTACGGGNGGCWGCAG) [10]. The amplified PCR products were bead purified and subsequent libraries were prepared using the Nextera, Illumina library prep kit (Cat. No. FC-131-1096). The quality of the library was assessed using a 2100 Bioanalyzer with a high sensitivity assay kit (Agilent CA, USA). The libraries were pooled in a reagent cartridge with standard single lane flow cell and were further sequenced on Miseq v3 system with 2 x 300bp paired-end chemistry.

### 2.4. Resistome analysis

The raw reads were analysed using the Microbial Genomics Module (CLC-MGM) in CLC Genomics Workbench 21.0.5 (Qiagen Bioinformatics, CA, USA) (Supplementary Fig.1).

Briefly, the raw reads were imported onto the CLC-genomics workbench, adapters were trimmed using the trim adapter tool. (Supplementary Table 2 for trimming statistics). The trimmed reads were aligned to the QMI-AR Peptide Marker Database to identify ARGs using DIAMOND v0.9.31 BLAST [11], under the Find Resistance with ShortBRED (Short Better Representative Extract Dataset), (FRSB) tool (similar to ShortBRED) with default settings (DIAMOND parameters: Genetic code-11 bacterial, archeal and plant plastid, E-value-0.00001, Sensitivity more sensitive search; Quantification parameters: Percent identity-0.95, Minimum alignment length-0.95, Minimum read length-90.0). FRSB tool maps the reads against a peptide database using a translational search, reports high-quality hits and normalises these count/abundances using maker length and sequencing depth [12]. It generates sample wise ARG abundance and resistance mechanism tables. The ARG abundance tables summarises the counts for each peptide marker is identified in each sample. The tables report normalised abundances as reads per kilo base per million mapped reads (RPKM) calculated based on marker length and sequencing depth as done in ShortBRED [12]. Peptide markers were further aggregated to calculate the abundance for individual ARGs using the aggregate function (Supplementary Table-4 & 5). The aggregated files were used for further analysis and ARGs were cross verified in the Comprehensive Antimicrobial Resistance Database (CARD) [13,14] for association with *ESKAPE* pathogens (*Enterococcus faecium, Staphylococcus aureus, Klebsiella pneumoniae, Acinetobacter baumannii, Pseudomonas aeruginosa, and Enterobacter species*) and other microbes.

### 2.5. Characterisation of the bacterial community

The raw reads from shotgun sequencing were analysed using the Microbial Genomics Module (CLC-MGM) in CLC Genomics Workbench 21.0.5 (Qiagen Bioinformatics, CA, USA) (Supplementary Fig. 2).

Trimmed shotgun sequencing reads were subjected to the taxonomic profiling tool on CLC-genomics workbench retaining the default settings. The reads were mapped against the QMI_PT Database [15]. This database represents all genera with variable number of species per genera. The Genome sequences and annotations are from the NCBI Reference Sequence Database.

Trimmed 16S RNA reads were subjected to OTU (Operational Taxonomic Unit) clustering tool under the Amplicon based taxonomic profiling tool in the microbial genomics module, with the default parameters for analysis. The reads were mapped using 97% similarity for OTU clustering using MiDAS (Microbial database for Activated Sludge) [16].

### 2.6. Statistical Analysis

All analyses were conducted using R v.3.4.2 [17]. First, the ARGs and taxa with more than 1% relative abundance were visualised using *ggplot2* package [18]. We used measures of alpha diversity (within-sample) indices (observed and Shannon index) in ARGs and microbial community to understand the differences sequencing methods. Alpha diversity indices were calculated using *vegan* package [19]. Wilcoxon tests were performed to test the statistical significance between methods and sample types.

In addition, we measured beta diversity (resistome composition) by sequencing methods. We performed Principal Coordinate Analysis (PCoA) using Bray-Curtis dissimilarity matrix, abundance weighted approach. Beta diversity was computed using *vegan* package. Permutational analysis of multivariate variance (PERMANOVA) was performed to test for statistical similarity across methods and sample types using Bray-Curtis distances. PERMANOVA was performed using the *vegan* package.

Spearman correlation was performed across methods, to understand the relationship between different sequencing methods using the *corr* function in R. The generated correlation coefficients (ρ) and p-values were visualised using *ggcorrplot* package [20].

Finally, we conducted a differential abundance analysis to estimate change in ARG abundance (>1%) from inlet to outlet samples by sequencing methods. The differential analysis was performed using the Differential Abundance Analysis tool under the Abundance analysis on CLC-genomics workbench. ARGs with log2 Fold change > 1 and p-values < 0.05 were considered were considered upregulated and the ARGs with log2 fold change < 1 and p-value<0.05 were downregulated.

## 3. Results and Discussion

### 3.1. Abundance, composition, and diversity of ARGs

Using three sequencing methods, a total of 785 ARGs were identified. The number of ARGs (761 ARGs) detected in inlet samples was much higher than in outlet samples (715 ARGs) and was consistent across sequencing methods (Fig. 1A, Table 1). Major ARGs, including aminoglycoside, beta-lactamase, fluoroquinolones, MLSB and tetracycline resistance genes were detected in all the samples. The number of ARGs detected in each sample ranged from 96 to 701, and the most abundant ARGs were found in inlet samples from KC1 STP across all the three sequencing methods.

**Figure 1:**
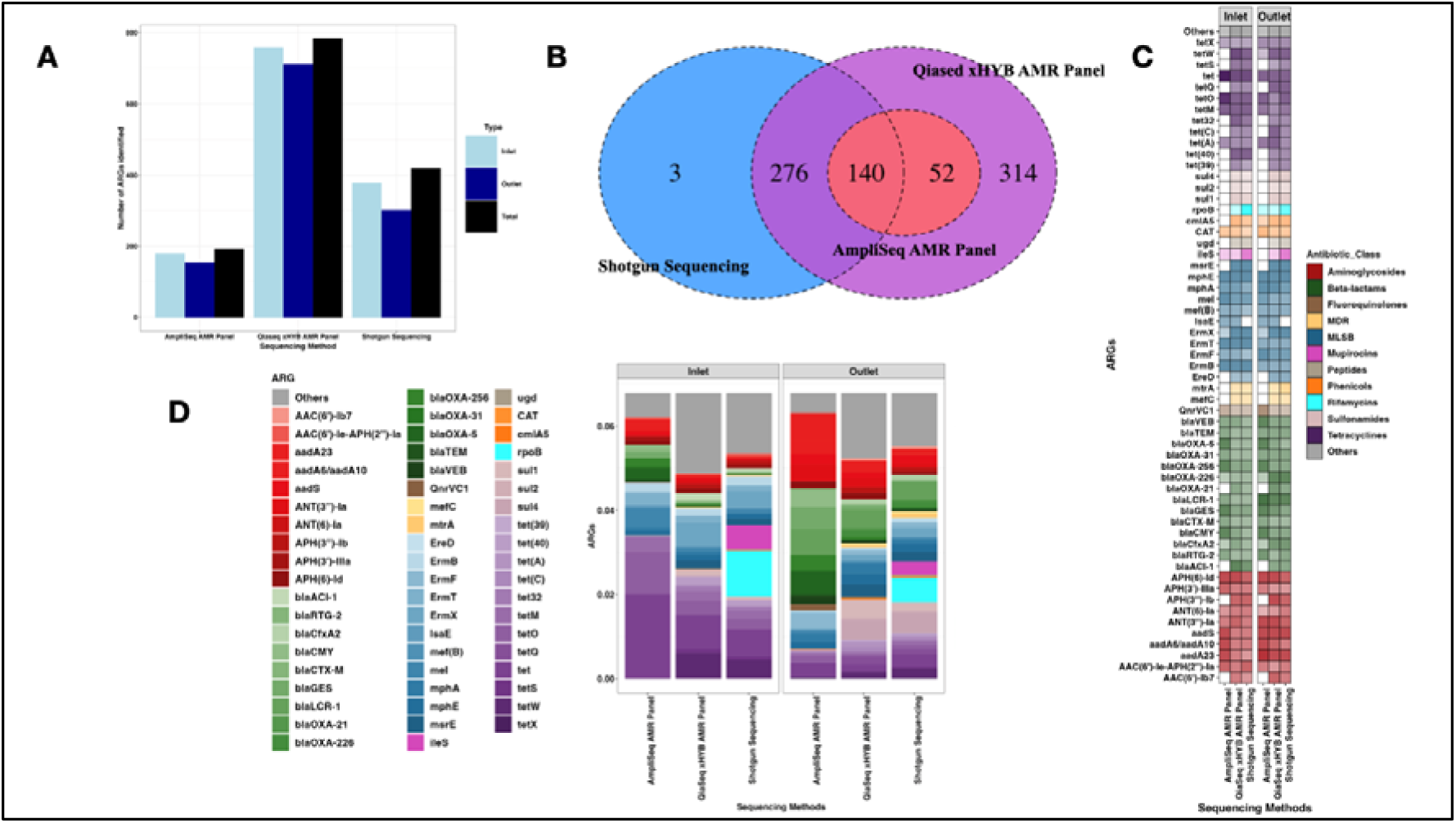
Comparison of ARGs detected by sequencing method and sample type. A) Abundance of identified ARG markers, quantified as reads per kilobase million (RPKM); B) Venn diagram showing the number of overlapping ARGs. C) Presence-absence heatmap showing the presence and absence of select ARGs (with relative abundance > 1%); D) Relative abundance of dominant ARGs (> 1% abundance). The relative abundances of all other ARGs less than 1% are grouped as “other.”

The QIAseq xHYB AMR panel identified the highest number of ARGs, 783 ARGs (759 in the inlet samples and 712 in the outlet samples) followed by shotgun sequencing with a total of 419 ARGs (378 in the inlet samples and 301 in the outlet samples). This heightened sensitivity of this panel could be attributed to an additional 16-hour hybridization target enrichment step in the library preparation, enhancing the detection of target sequences [13]. In contrast, the AmpliSeq AMR panel identified lowest number of ARGs with a total of 192 (179 in the inlet and 155 in the outlet).

Out of the 783 ARGs, we observed 140 (18.47%) ARG types overlapped across sequencing methods (Fig. 1B). In contrast, 314 ARGs (40%) were exclusively detected using QIAseq xHYB AMRpanel and 3 ARGs (0.38%) were exclusively identified by shotgun sequencing. The AmpliSeq panel showed complete overlap in ARGs types with the QIAseq xHYB panel. In contrast, AmpliSeq panel showed only ARGs overlapping with shotgun sequencing. QIAseq xHYB AMR panel and shotgun sequencing showed an overlap of 416 ARGs (52.9%).

Despite differences in the number of ARGs detected, the overall diversity remained statistically non-significant across methods. Our pairwise correlation analysis of overlapping ARGs revealed QIAseq xHYB AMR panel and shotgun sequencing showed high correlation in terms of relative abundances (ρ =0.88), whereas AmpliSeq AMR panel showed relatively weaker correlations with QIAseq xHYB AMR panel (ρ = 0.65) and shotgun sequencing (ρ =0.59), indicating that the methods are comparable in terms of efficiency to detect ARGs (Supplementary Fig. 3).

Based on the proportions of ARGs detected, tetracycline (24.8%), MLSB (17.33%), beta-lactams (15.04%), and aminoglycosides (12.41%) were four dominant ARG antibiotic classes detected by each sequencing method. However, there varying patterns in ARG composition by sample type and sequencing method. For example, Rifamycins (*rpoB*) and Mupirocins (*ileS*) were only detected by shotgun sequencing method in the inlet and outlet samples. The AmpliSeq AMR panel identified *tetO* (14.75%) and *aadA23* (8.9%) as the most abundant ARGs in the inlet and outlet, respectively. The QIAseq xHYB AMR panel and shotgun sequencing identified *tetW* (9%) *rpoB* (16%) as the most dominant ARGs in the inlet respectively. In the outlet *sul4* was identified as the most abundant ARG by both QIAseq xHYB AMR panel (6.87%) and shotgun sequencing (7.5%). *blaCTX-M,* commonly associated with *E. coli* and *tetX* associated with urinary tract infection causing bacteria belonging to *Enterobacteriaceae* and *Pseudomonadaceae,* were identified as dominant ARGs by all the three sequencing methods [21].

The presence of ARGs in sulfonamide antibiotic class such as *sul1* in sewage implicate strong anthropogenic contamination and increased rates of horizontal gene transfer (HGT) [22]. QIAseq xHYB AMR panel and shotgun sequencing were the only methods that identified *sul1, sul2 and sul4* as dominant ARGs. Similarly *tet(40),* commonly associated with the gut microbiome was identified by QIAseq xHYB AMR panel and shotgun sequencing as a dominant ARG. Similarly, clinically relevant carbapenems ARGs *blaNDM-1, blaVIM, blaKPC and blaIMP* were only detected by QIAseq xHYB AMR panel, albeit in low abundance (< 0.00001). Similar results were also observed in a study in Japan [22].

Colistins are used as the last line of defence against infections caused by carbapenem-resistant bacteria. Colistin resistance conferred by *mcr-1, mcr-3* is often observed in clinical isolated of *Escherichia coli* and *Klebsiella pneumoniae* making these ARGs of key importance [23]. QIAseq xHYB AMR panel identified *mcr-1* ARG along with other members of *mcr* gene family. Shotgun sequencing was unable to identify *mcr-1* but was able to identify other members of the *mcr* gene family (*mcr-8*, *mcr-9),* whereas AmpliSeq AMR panel failed to identify members of *mcr* gene family. *blaTEM,* conferring resistance to penicillin, cephalosporins and carbapenems is one of the most common ARG that is known to be transferred by plasmids. These antibiotics account for approximately 2/3^rd^ of the total antibiotic weight consumed in the world [24, 25]. *blaTEM* was detected across all the three methods with an increased abundance in the outlet. *aadA* family of ARGs confers resistance to streptomycin’s that are often used as the first line of drugs for tuberculosis treatments [26]. Members of *aadA* ARG family such as *aadA23, aadA6/aadA10* were identified across sequencing methods in variable abundances.

Alpha diversity measures observed richness and Shannon diversity showed no significant difference (*p>*0.50) by sample type or sequencing method (Figure 2A and 2B). The PCoA showed distinct clusters by inlet and outlet samples across three sequencing methods (Figure 2C). PERMANOVA analysis revealed that the differences were statistically significant by sequencing method (inlet: R^2^=0.93, *p*<0.001, outlet: R^2^=0.80, *p*<0.001).

**Figure 2:**
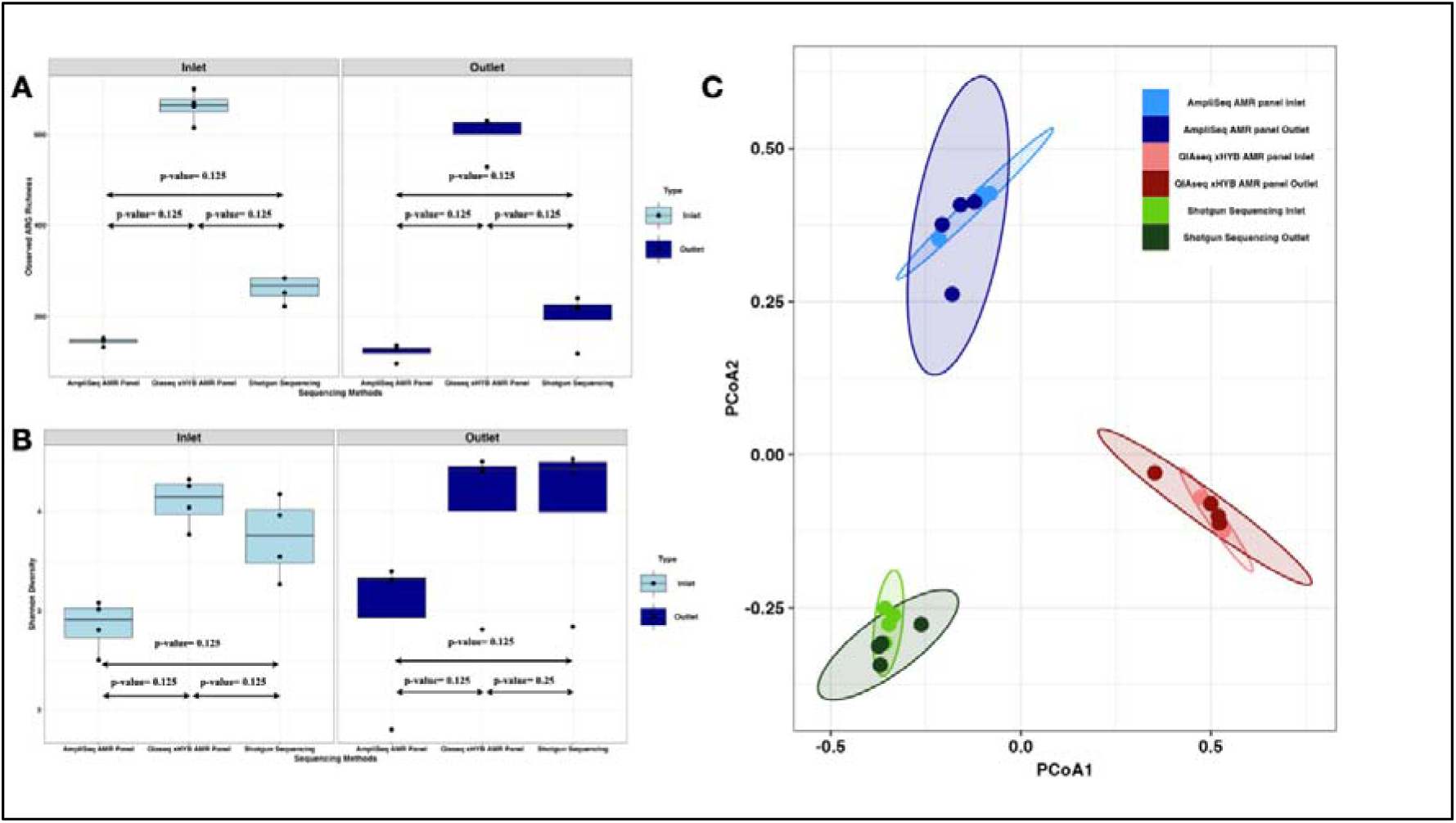
Alpha and Beta diversity measures by sample type and sequencing methods. A) Observed richness, B) Shannon diversity, C) PCoA.

To explore shifts in ARG abundance from inlet to outlet, we used 59 dominant ARGs (>1% abundance) shared across all sequencing methods (AmpliSeq panel: 32, QIAseq xHYB AMR panel: 41, shotgun sequencing: 45). For example, beta-lactams (*blaLCR-1*, *blaOXA-226*) and sulphonamides (*sul4*) showed significant increase in value in outlet samples whereas tetracycline (*tet* (40), *tetS*, *tetO*) showed significant decrease in their abundances in outlet samples (Figure 3). *blaLCR-1* showed significant increase in value across three methods (Ampliseq AMR panel: *FC_log2_* =7.15; QIAseq xHYB AMR panel: *FC_log2_*=7.76; Shotgun sequencing: *FC_log2_*=6.36). *APH (3’)-IIIa* showed significant decrease in its abundance (Ampliseq AMR panel: *FC_log2_*=-3.58, QIAseq xHYB AMR panel: *FC_log2_*= −2.56). *ErmB* showed significant decrease in value in shotgun sequencing (*FC_log2_*=−3.46). *Mef(B)* and *sul1* showed a decrease in value in outlet samples in AmpliSeq AMR Panel, and an increase in abundance values in the shotgun sequencing and QIAseq xHYB AMR Panel (Figure 3; Supplementary table 3)).

**Figure 3:**
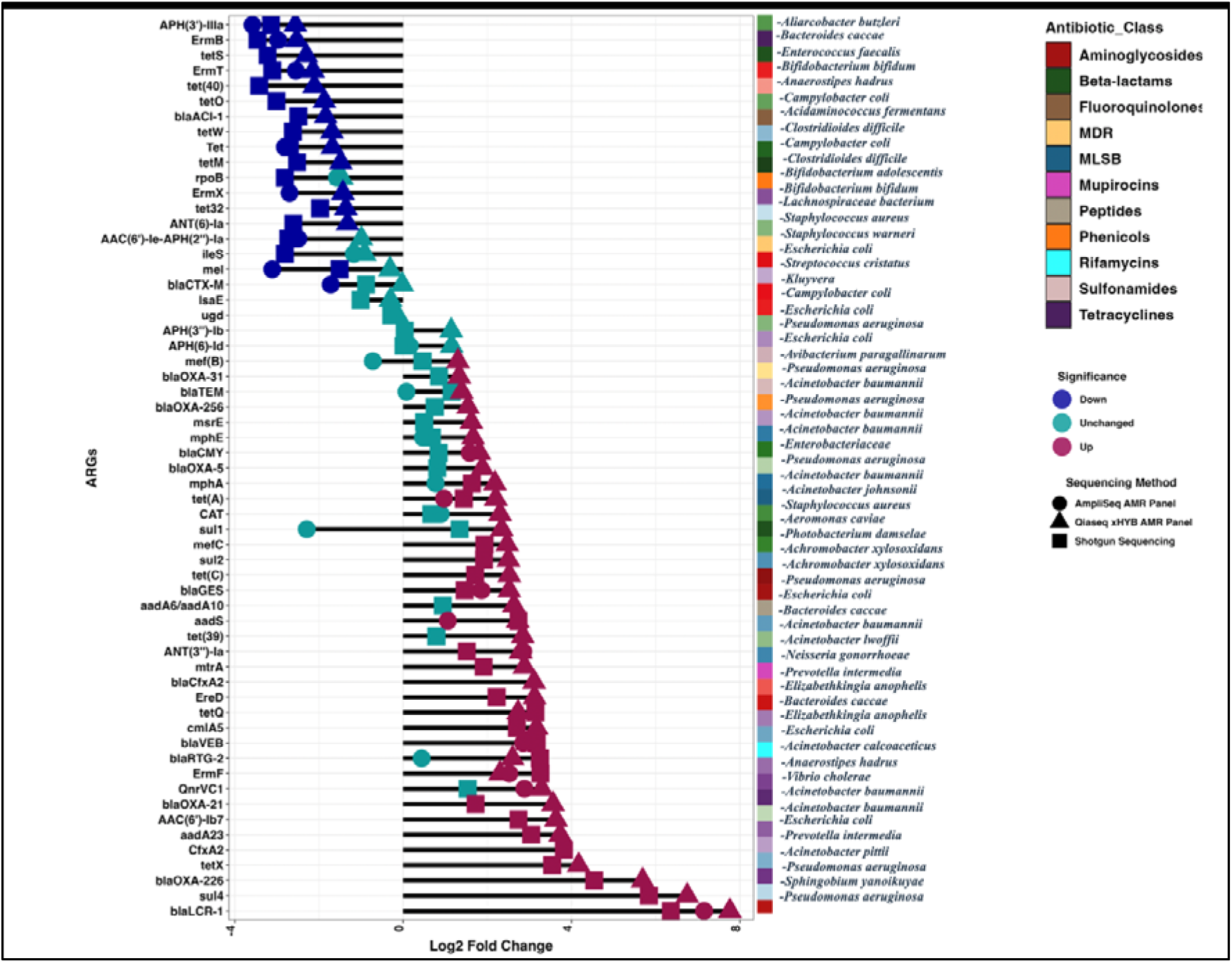
Differential abundance analysis exploring shifts in ARGs abundance from inlet to outlet samples. Green coloured icons represent non-significant (*p* >0.05) referred as unchanged, blue (down) and magenta (up) icons indicate significant reduction and increase in abundance of ARGs (*p*-values <0.05) in the treated samples with respect to non-treated samples, respectively. The microbes associated with the enlisted ARGs based on the resistome matches from CARD database have been marked on the y-axis.

ARGs associated with *ESKAPE* pathogens *blaLCR-1, blaGES, blaOXA-226* associated with *Pseudomonas aeruginosa* showed significant increase in abundance in the outlet samples. *CAT* associated with *Staphylococcus aureus* showed significant increase only in QIAseq xHYB AMR panel. *AAC (6’)-Ie-APH2-Ia* associated with *Staphylococcus* species, showed significant decrease in abundance in AmpliSeq and QIAseq xHYB AMR panels.

### 3.2. Bacterial taxonomy using shotgun metagenomic sequencing and 16S RNA amplicon-based sequencing

Both sequencing methods identified *Acinetobacter* and *Firmicutes* as the most abundant phyla in the inlet samples. Whereas *Proteobacteria* and *Bacteroidota* were the most abundant phyla in the outlet samples (Fig.4 A and B).

**Figure 4:**
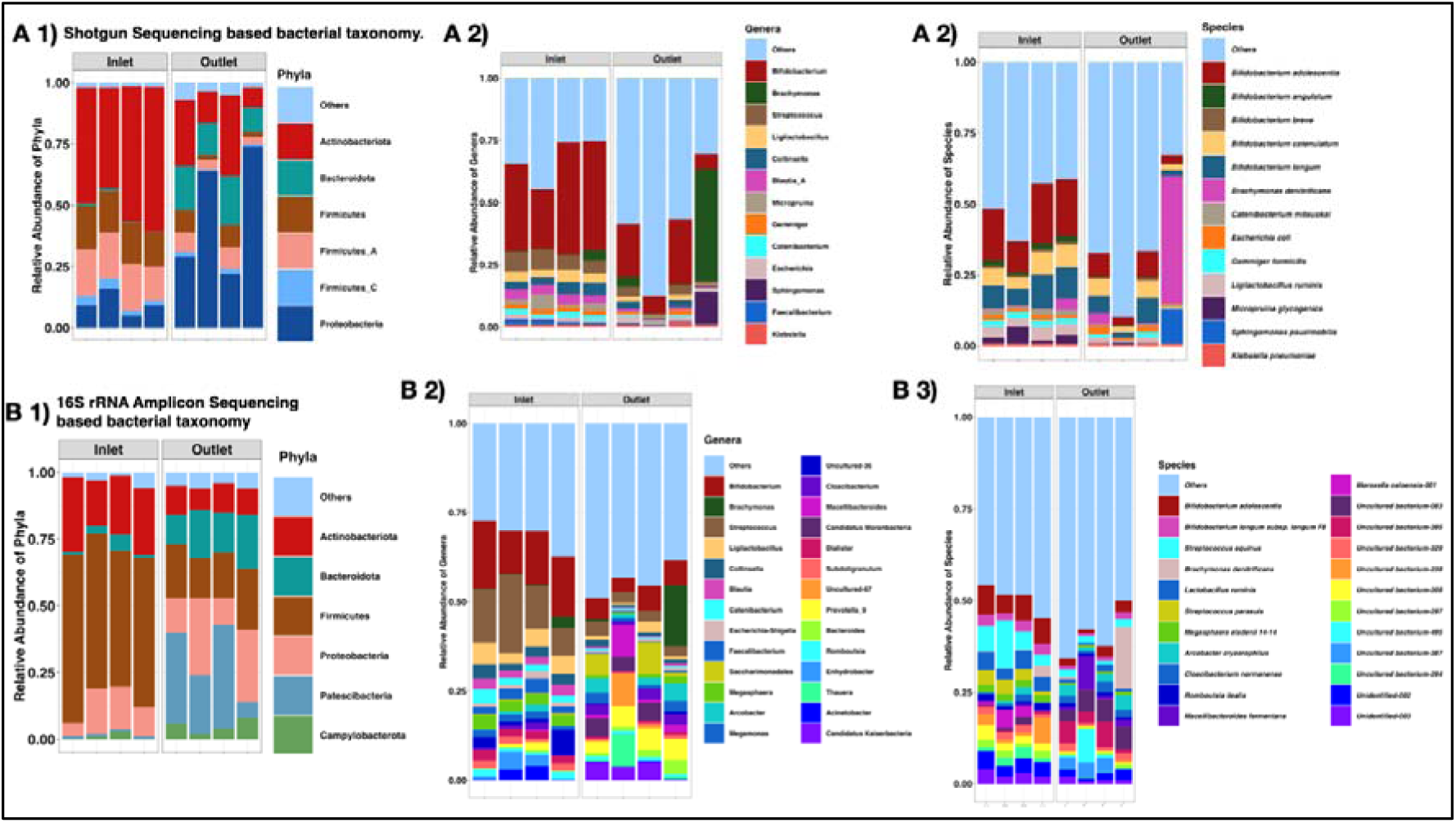
Bacterial taxonomic profile of phyla, genera and species with greater than 1% abundance of inlet and outlet samples using shotgun sequencing and 16S Amplicon-based sequencing. A) Abundance plot for dominant phyla for shotgun sequencing. B) Alpha diversity indices (observed richness and Shannon Index) across inlet and outlet samples for shotgun sequencing. C) Abundance plot for dominant phyla for 16S amplicon-based sequencing. D) Alpha diversity indices (observed richness and Shannon Index) across inlet and outlet samples for 16S amplicon-based sequencing.

At the genera level, both sequencing methods identified *Bifidobacterium* among the top 10 genera in the inlet and outlet samples. However, *Streptococcus* and *Brachymonas* were the abundant phyla in the outlet samples. 16S RNA amplicon-based sequencing identified, *Streptococcus equinus* and *Brachymonas denitrificans* as the most abundant species in the inlet and outlet sewage samples respectively, whereas shotgun sequencing identified *Bifidobacterium adolescentis* and *Brachymonas denitrificans* as the most abundant species in the inlet and outlet respectively. Shotgun sequencing detected the *ESKAPE* pathogen family (*Klebsiella pneumoniae, Escherichia coli)* as dominant species (with relative abundance greater than 1%), however, 16S rRNA based amplicon sequencing was unable to capture these as dominant species. 16S rRNA sequencing identified, many uncultured and unidentified bacteria as these might have not been annotated in the database used for BLAST search that were not observed in the shotgun sequencing.

Alpha diversity indices were not significantly different by sequencing methods and sample types (Fig. 5 A and B).

**Figure 5:**
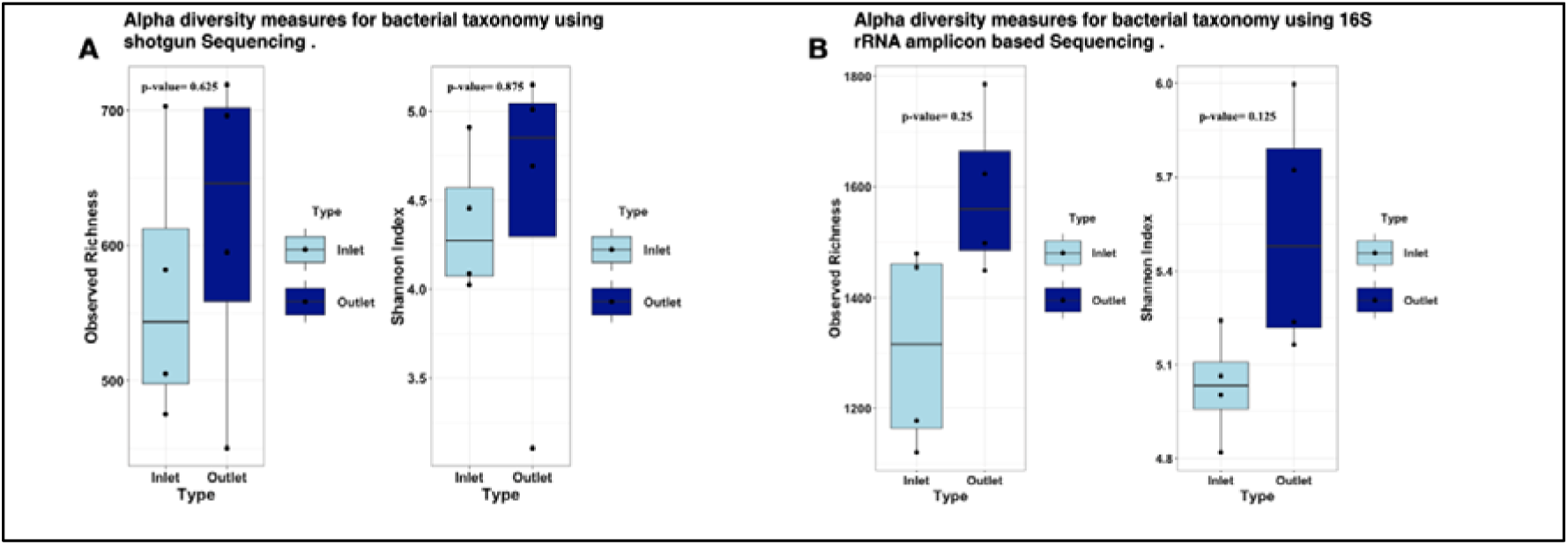
Alpha diversity indices for bacterial taxonomy using shotgun sequencing and 16S rRNA amplicon-based sequencing. A) Observed richness and shannon diversity indices for bacterial taxonomy using shotgun sequencing. B) Observed and Shannon diversity indices for bacterial taxonomy using 16S rRNA based amplicon sequencing.

Whilst the cost-effectiveness analysis revealed comparable per-sample costs for all three platforms, it is important to highlight that amplicon-based methods (Ampliseq for AMR Panel and Qiaseq xHYB AMR Panel) targeted a specific number of genes (478 ARGs and 2786 ARGs, respectively) which may or may not be present in the given sample, in contrast with shotgun sequencing considers large whole genome approach amplifying all nucleic acid content, offering a broader scope of information, including bacterial as well as nonbacterial taxonomy. This highlights a trade-off between specificity and inclusivity, with amplicon-based platforms enriching only for ARGs and lacking additional taxonomic information. While the QIAseq xHYB AMR Panel demonstrates enhanced sensitivity [27].

In the differential analysis of dominant ARGs observed consistent trends across methods, with ARGs showing similar changes in abundance levels from the inlet to the outlet, although the degree of change varied which could reflect in gauging the efficiency of sewage treatment plant. The Qiagen AMR panel exhibited a significant increase in ARG abundance for all identified genes in the outlet compared to the inlet across sequencing methods. Nonetheless, all three methods provide valuable insights into for the prevalence and diversity of ARGs in sewage and should be considered in the context of the study. The dominant ARGs identified using these sequencing methods can be selectively targeted for developing surveillance panels or RT-PCR based quantification as these ARGs are implicated to be associated with most of the high priority *ESKAPE* pathogens and can provide indications about increased rates of transmission by HGT. Performances of sequencing methods have shown dependence on several factors including sample size [28]. With this in mind, researchers and practitioners must consider the specific goals and trade-offs associated with each method when selecting a suitable tool for AMR surveillance in each setting.

## Data Availability

All data produced in the present study are available upon reasonable request to the authors.

## Acknowledgments

We sincerely thank Bengaluru Water Supply and Sewerage Board for providing access to the sewershed sites. We thank Dr. Vishwanath, for providing us with detailed understanding of the sewage network in Bengaluru. We are grateful to NGS facility at National Centre for Biological Sciences for facilitating the sequencing of wastewater samples. We thank Annamalai N for sample processing and library preparation.

## Author’s contribution

DM: CLC-genomics analysis, visualisation using R and writing the first draft of the manuscript.

FI: Conceptualised the project, supervised, analysis and writing and editing the draft.

## Funding

This work has been supported by the funding from Tata Trust to Tata Institute of Genetics and Society (TIGS) and Rockefeller foundation (grant number 2021 HTH 018).

## Conflict of interest

The authors declare that they have no conflict of interest.

## Ethical approval

Not required

---

**Supplementary Figure 1:**
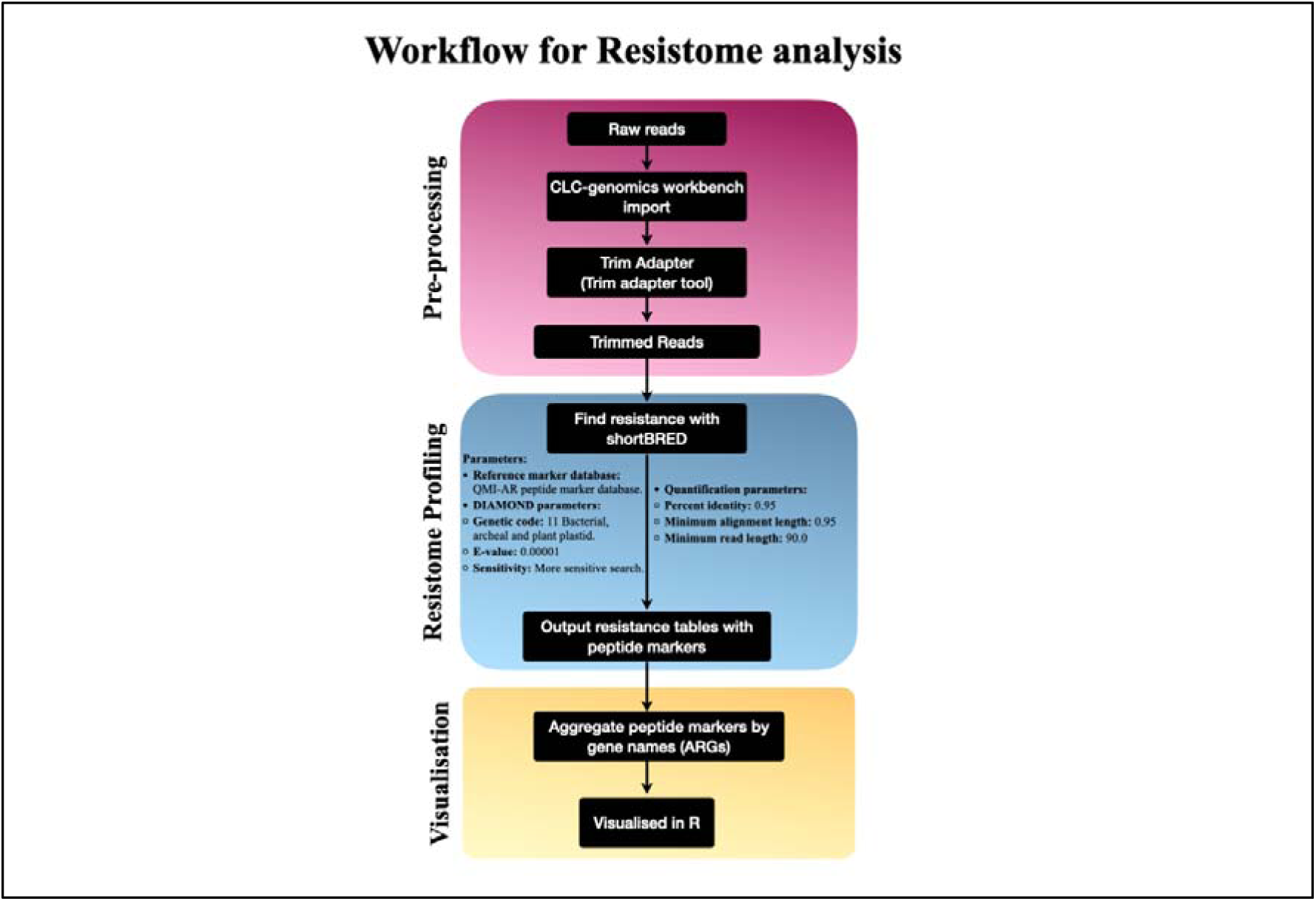
CLC-Genomics workflow for resistome analysis.

**Supplementary figure 2:**
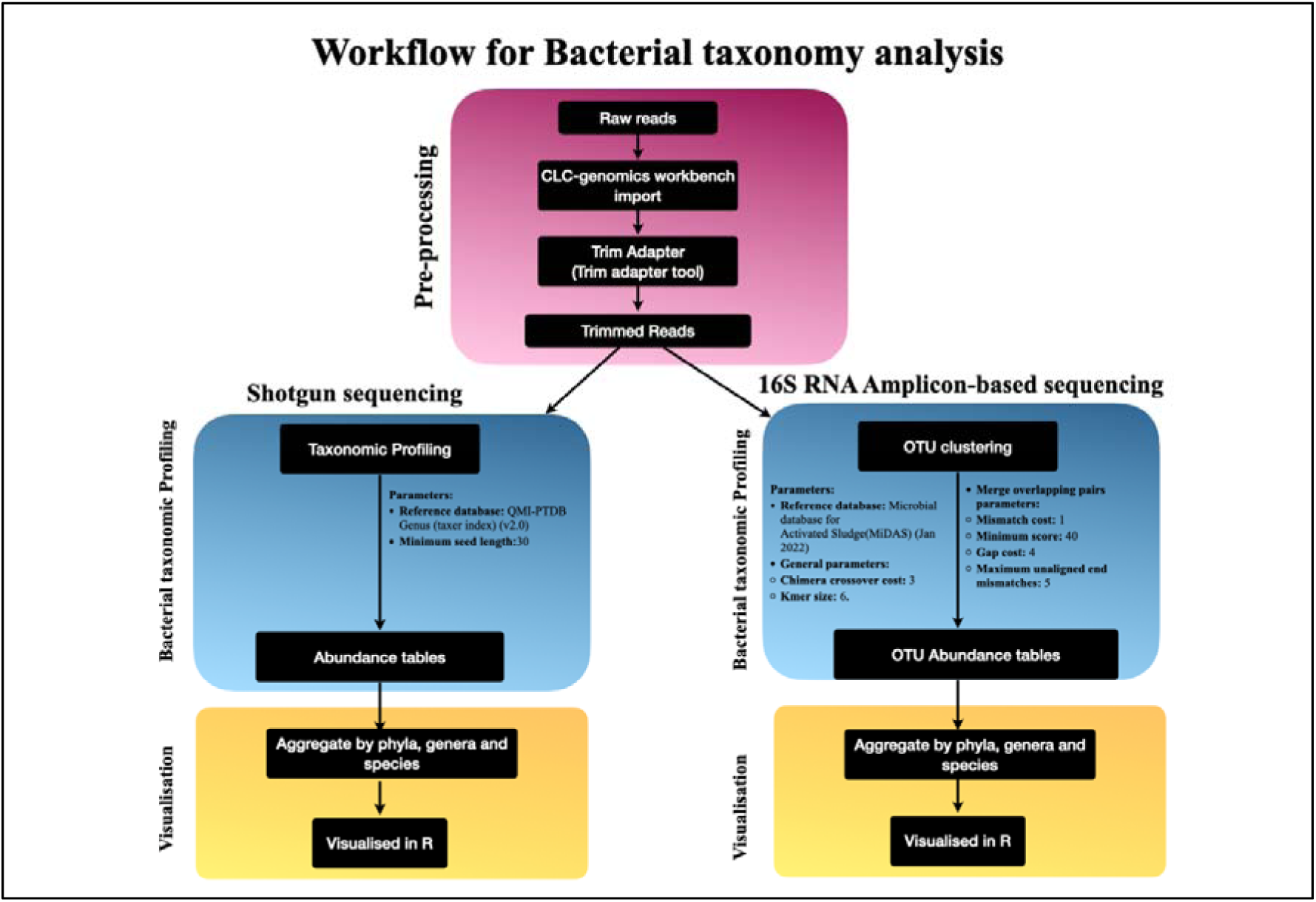
Bacterial taxonomic profiling workflow CLC-Genomics

**Supplementary Figure 3:**
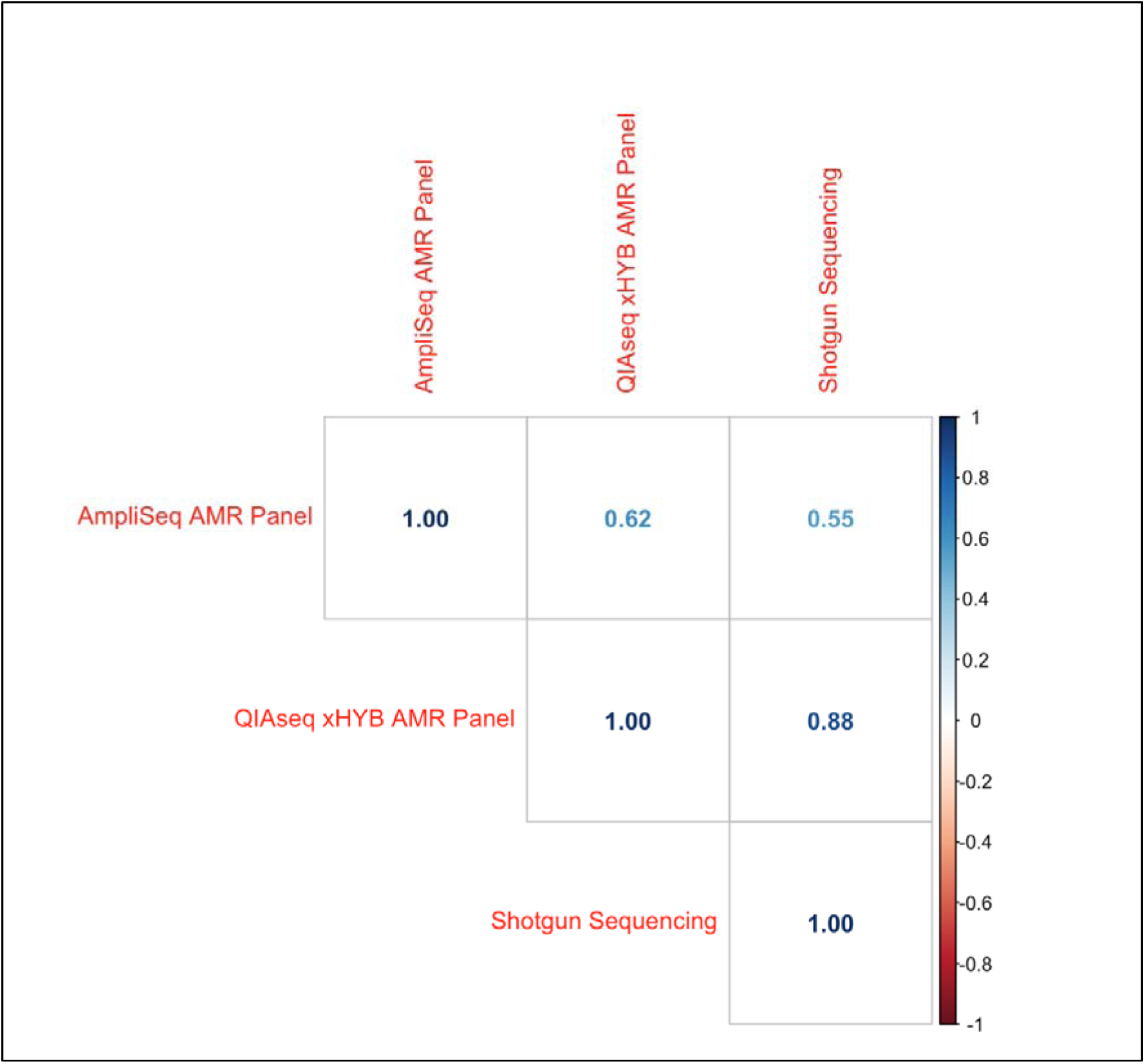
Spearman correlation between 140 overlapping ARGs across sequencing methods

## Notes

### Competing Interest Statement

The authors have declared no competing interest.

